# Predicting vaccine hesitancy from area-level indicators: A machine learning approach

**DOI:** 10.1101/2021.03.08.21253109

**Authors:** Vincenzo Carrieri, Raffele Lagravinese, Giuliano Resce

## Abstract

Vaccine hesitancy (VH) might represent a serious threat to the next COVID-19 mass immunization campaign. We use machine-learning algorithms to predict communities at a high risk of VH relying on area-level indicators easily available to policymakers. We illustrate our approach on data from child immunization campaigns for seven non-mandatory vaccines carried out in 6408 Italian municipalities in 2016. A battery of machine learning models is compared in terms of area under the Receiver Operating Characteristics (ROC) curve. We find that the Random Forest algorithm best predicts areas with a high risk of VH improving the unpredictable baseline level by 24% in terms of accuracy. Among the area-level indicators, the proportion of waste recycling and the employment rate are found to be the most powerful predictors of high VH. This can support policy makers to target area-level provaccine awareness campaigns.

## 1. Introduction

Vaccine hesitancy (VH) may constitute a serious threat to the mass COVID-19 immunization campaign. Opinion polls in France and US on vaccination intentions suggest that COVID-19 vaccine hesitancy is increasing worldwide (Schwarzinger et al. 2021; Reiter et al. 2020). A recent survey indicates potential acceptance rates for COVID-19 vaccines ranging from 55% to 90% (Lazarus et al. 2020). Beyond COVID-19, VH represents a threat also to the eradication of re-emergent vaccine-preventable diseases like measles and rubella (Horne et al. 2016; WHO, 2019; ECDPC, 2019).

Evidence has accumulated that several individual-level factors are strongly associated with VH, such as low income and poor education, certain political or religious orientations, low trust in conventional medicine and institutions, and risk perception (Dincer and Gillanders 2021; Caserotti et al. 2021; Buckman et al., 2020; Hornsey et al., 2020; Ward et al., 2020; Chang, 2018; Motta et al. 2018; Yakub et al., 2014). However, in practice, health authorities typically do not have such detailed individual-level information available to them in sufficient time to be useful for informing immunization campaigns and/or can be very costly to design *ad-hoc* surveys for this purpose. Moreover, immunization campaigns are usually organized at the local level (typically municipality level). Thus, to target appropriate interventions, would be extremely useful for policy makers to know on the basis of area-level indicators easily available to them (institutional features, demographic and geographic factors, and socio-economic indicators), the communities more at risk of HV and identify the set of area-level correlates of VH. This is particularly important also to enforce local herd immunity and limit the spreading of a disease to other areas of a country.

In this paper we propose a practical approach to identify “hotspots” for VH relying on lagged municipality-level indicators easily available to policy-makers and machine-learning algorithms. The application of machine learning techniques to health issues has increased in the recent times. Applications range from the relationship between height and socio-economic status (Daoud et al. 2019) to socio-economic determinants of health (Seligman et al. 2018) and key correlates of child marriage (Raj et al. 2020). A few papers also employed machine learning algorithms to predict VH using patient-level data. For instance, Bell et al (2019) use machine learning approach to identify children at risk of not being vaccinated against Measles, Mumps and Rubella in WHO countries. Oreskovic et al. (2020) adopt Proactive machine-learning-based approaches to VH for a potential SARS-Cov-2 using LASSO logistic regression on a low number of attributes of the child and his or her family and community.

The machine-learning approach offers several policy-relevant advantages in the analysis of VH using area-level indicators. First, it allows us to overcome the problem of incomplete data on immunization coverage which represents a key issue particularly in low- and middle-income countries (Harrison et al. 2020). Second, it allows us to predict areas at high VH risk relying on coverage data from previous immunization campaigns. This might help to get valuable insights also on the local acceptance rates for the next COVID-19 immunization campaign. Last but not least, the identification of the most powerful predictors of VH allows us also to derive a parsimonious set of indicators that can be employed in cases in which only a small set of area-level indicators is available. Again, this is of particular relevance in areas in which data collection is challenging.

We illustrate our approach using real-life data from the child immunization campaigns for seven vaccine-preventable carried out in 6408 municipalities in 2016 in Italy. We focus on child immunization relative to the babies turning 24 months. We only consider the seven vaccines that until 2016 were recommended but not mandatory^1^. Italy represents an interesting setting to study VH because of the high prevalence of no-vax feelings. The spreading of fake news on the web (Carrieri et al. 2019) and the high support to political parties repeatedly campaigned on an anti-vaccination platform (i.e., The Five-Star Movement - M5S) correlated with a recent spread of vaccine-preventable diseases. For instance, among European countries, Italy (4,978 cases) had the highest incidence of measles cases between 1 February 2017 and 31 January 2018 (ECDC, 2019).

We find that the Random Forest algorithm is the best model to make out-of-sample predictions of areas with high VH, with a high true positive rate and low false-positive rate. The final model predicts with high accuracy (0.761), sensitivity (0.623), and specificity (0.850). Compared with the unpredictable baseline level, the accuracy with Random Forest improves by 24%. Remarkably, our best model shows performances in line with recent studies that make use of patient level indicators with a higher granularity (see Bell et al. 2019). Among the area-level indicators, the share of waste recycling and the employment rate are found to be the most powerful predictors. This offers strong policy implications for the next COVID-19 immunization campaign.

The rest of paper is structured as follows. Next section presents data and methodology. Section 3 shows results. Last section summarizes and concludes.

## 2. Data and Methodology

Table 1 shows a set of municipality-level indicators collected by the Italian National Institute of Statistics (ISTAT) and the Italian Ministry of the Interior. These data are matched with data collected by the Italian Ministry of Health about child immunization campaigns done in 6408 Italian municipalities in 2016 for seven vaccine-preventable diseases (pertussis, measles, haemophilus influenzae type B, meningococcus, pneumococcus, mumps, and rubella).

**Table 1.** Municipal cha0racteristics

| Demographics and geographical ($D_x^t$) | Institutional ($I_x^t$) | Socio-economic ($S_x^t$) |
| --- | --- | --- |
| Population Density | Proportion of recycling waste | Employment rate |
| Altitude | Average age of the council's members | Proportion of temporary jobs |
| Divorce rate | Age of the Mayor | Education |
| Proportion of families with Child | Proportion of females in the Council | Poverty |
| Population | Special Statute Regions | Average Income |
| Proportion of population under 18 | Mayor of Centre-Left | Inequality |
| Proportion of population over 65 | Southern Region | Access to Internet |
| Proportion of population under 2 | Mayor of Centre-Right |  |
| Internal Area | Mayor with the degree or more |  |
| Earthquake | Female Mayor |  |
|  | Municipal Default |  |

Each municipality *x* at year *t* consists of the following group of characteristics: Demographics and geographical 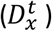, Institutional 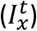, and Socio-economic 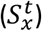.

Every municipality *x* at time *t* has an associated target binary variable 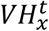 (vaccine hesitancy) that takes values “1” (positive sample) if the average child vaccine coverage is under 92%^2^, and value “0” (negative sample) otherwise. The prediction task is formulated as follows: based on the set of features 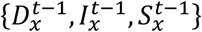 for municipality *x* find function *f*(.) (Machine Learning model) that predicts vaccine hesitancy 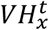:

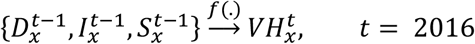

We randomly divide the database as 70 percent for training and 30 percent for out-of-sample testing set (test). The hyper-parameter optimization is only done on the training set. Four different models have been analysed: Least Absolute Shrinkage and Selection Operator (LASSO - Tibshirani, 1996), Random Forest (RF - Breiman, 2001), Neural Network (NN - Venables and Ripley, 2002) and Gradient Boosted Machine (GBM - Friedman, 2001).

The performance of vaccine hesitancy classification prediction is assessed by the Receiver Operating Characteristics (ROC) curve (Fawcett, 2006). In our binary classification problem, the positive class is defined as the municipality with high VH risk and the negative class is the municipality with low VH risk. The ROC curve shows the classifier diagnostic ability by plotting the true positive rate (TPR) on the *y*-axis against the false positive rate (FPR) on the *x*-axis since its discrimination threshold is varied (Antulov-Fantulin et al. 2021). The true positive rate is the ratio of municipalities with high VH risk that were correctly categorized as high VH risk (true positive) and the total number of positive samples (high VH risk). The false positive rate is the ratio between the number of municipalities with low VH risk wrongly categorized as high VH risk (false positives) and the total number of actual negative samples (low VH risk). When the classification task is completely unpredictable, the negative class theoretical distribution over feature space coincides with the positive class theoretical distribution over feature space. This implies that the ROC curve would be the diagonal line with an Area Under the Curve (AUC) of 0.5. A perfect classifier has AUC equal to the 1.0, the higher the AUC, the more predictive is the model.

Machine learning models also give information on how useful each feature is in explaining VH. Each model has a different algorithm to estimate importance. In LASSO, feature importance is estimated as the absolute value of the coefficients corresponding to the tuned model (Kuhn, 2020). For RF, feature importance is the mean gain produced by the feature over all the trees where the gain is measured by the Gini index (Liaw, Wiener, 2002). The feature importance in GBM is the average improvement of the splitting on the features across all the trees generated by the boosting algorithm (Friedman, 2001; Ridgeway, 2007). NN uses the Garson (1991) algorithm to evaluate relative variable importance. In this case the importance of each variable is determined by identifying all weighted connections between the layers in the network (Venables Ripley, 2002).

## 3. Results

Figure 1 shows the ROC curves for four different algorithms: LASSO, RF, NN, GBM. The estimates are based on the cross-validation algorithm which trains and tests the model tuning the hyper-parameters with the aim of maximising the area under the ROC curve.

**Figure 1.**
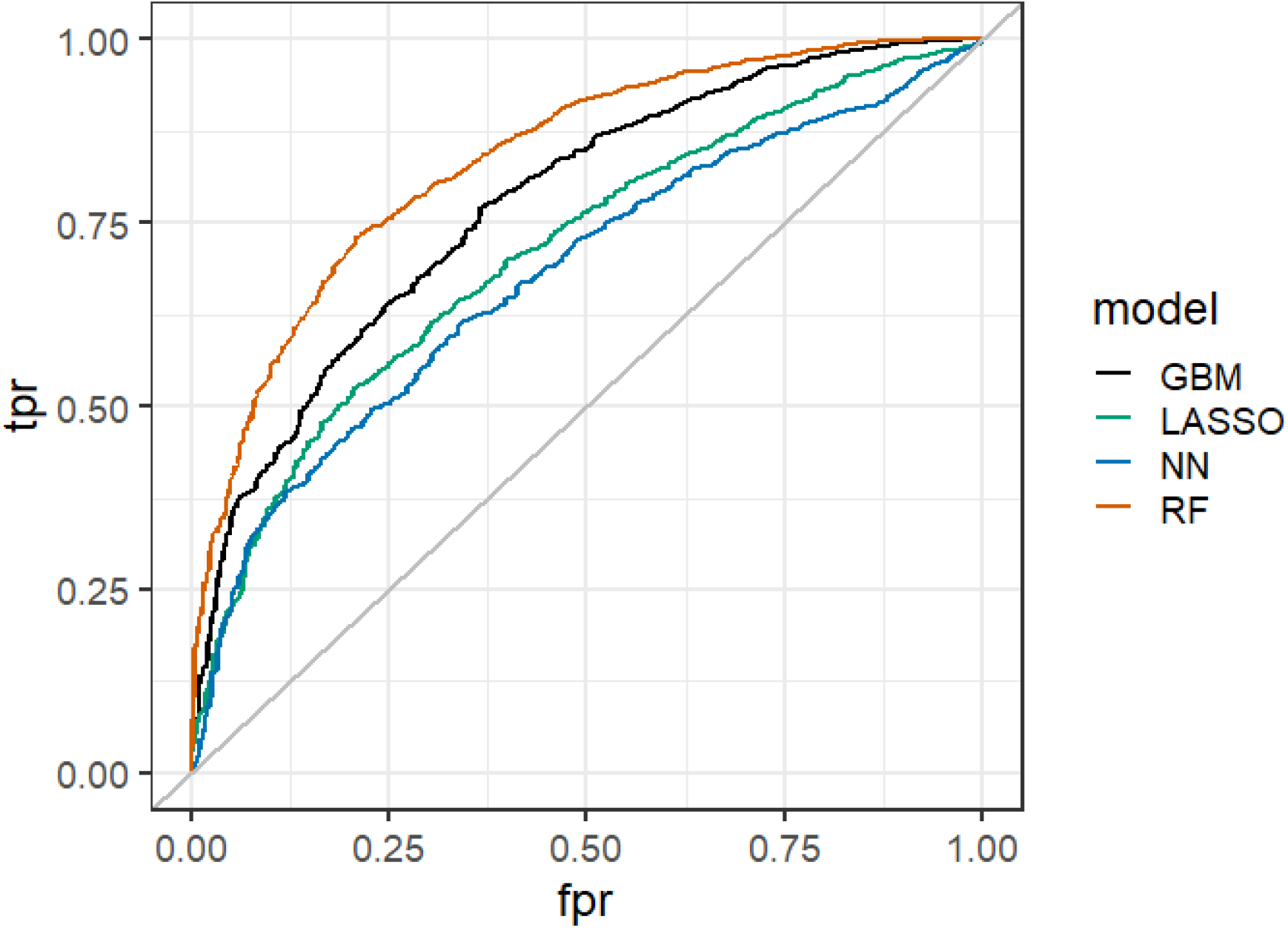
ROC curve for four Machine Learning models. Models trained on 70% of observations and tested on the remaining 30%. GBM (Gradient Boosted Machine) AUC = 0.775, LASSO (Least Absolute Shrinkage and Selection Operator) AUC = 0.709, NN (Neural Network) AUC = 0.677, and RF (Random Forest) AUC = 0.836. Resampling: Cross-Validated (10-fold, repeated 5 times)

The area under the ROC curve (AUC) in Figure 1 shows that the Random Forest model outperforms all the other models (AUC: RF = 0.836; GBM = 0.775; LASSO = 0.709; NN = 0.677). The best model has accuracy =0.761 (95 per cent CI:0.742, 0.782), sensitivity = 0.623, and specificity =0. 850 (see Table 2).

**Table 2.** Model’s performances

|  | **RF** | **GBM** | **NN** | **LASSO** |
| --- | --- | --- | --- | --- |
| Accuracy | 0.761 | 0.717 | 0.644 | 0.662 |
| 95% CI | (0.742, 0.782) | (0.695, 0.737) | (0.621, 0.665) | (0.640, 0.684) |
| No Information Rate | 0.614 | 0.614 | 0.614 | 0.614 |
| P-Value [Acc > NIR] | 0.000 | 0.000 | 0.006 | 0.000 |
| Sensitivity | 0.623 | 0.558 | 0.398 | 0.506 |
| Specificity | 0.850 | 0.817 | 0.798 | 0.760 |
| Pos Pred Value | 0.722 | 0.656 | 0.553 | 0.570 |
| Neg Pred Value | 0.782 | 0.746 | 0.679 | 0.711 |
| Prevalence | 0.386 | 0.386 | 0.386 | 0.386 |
| Detection Rate | 0.240 | 0.215 | 0.154 | 0.195 |
| Detection Prevalence | 0.333 | 0.328 | 0.278 | 0.343 |
| Balanced Accuracy | 0.737 | 0.687 | 0.598 | 0.633 |
| Area under the ROC Curve | 0.836 | 0.775 | 0.677 | 0.709 |

To test whether the differences among the ROC curves are significant, we use the DeLong’s statistical test (DeLong et al., 1988; Robin et al., 2011). Results in Table 3 show that the difference between the ROC curve of the Random Forest model and all the other models is statistically significant (p-value < 0.0005).

**Table 3:** Results of DeLong’s test for ROC curves

|  | **Z** | **p-value** |
| --- | --- | --- |
| RF vs GBM | 9.208 | 0.000 |
| RF vs LASSO | 12.056 | 0.000 |
| RF vs NN | 12.804 | 0.000 |

Figure 2 reports the first ten most relevant features for the Random Forest model. The share of recycling waste and employment rate are the top two area-level features predicting high VH risk. Among the top ten features associated with high VH risk there are also the altitude, poverty rate, the share of families with children, the share of temporary jobs, the divorce rate, the average education level (, i.e. the share of population with a degree or more), population density, and total population.

**Figure 2.**
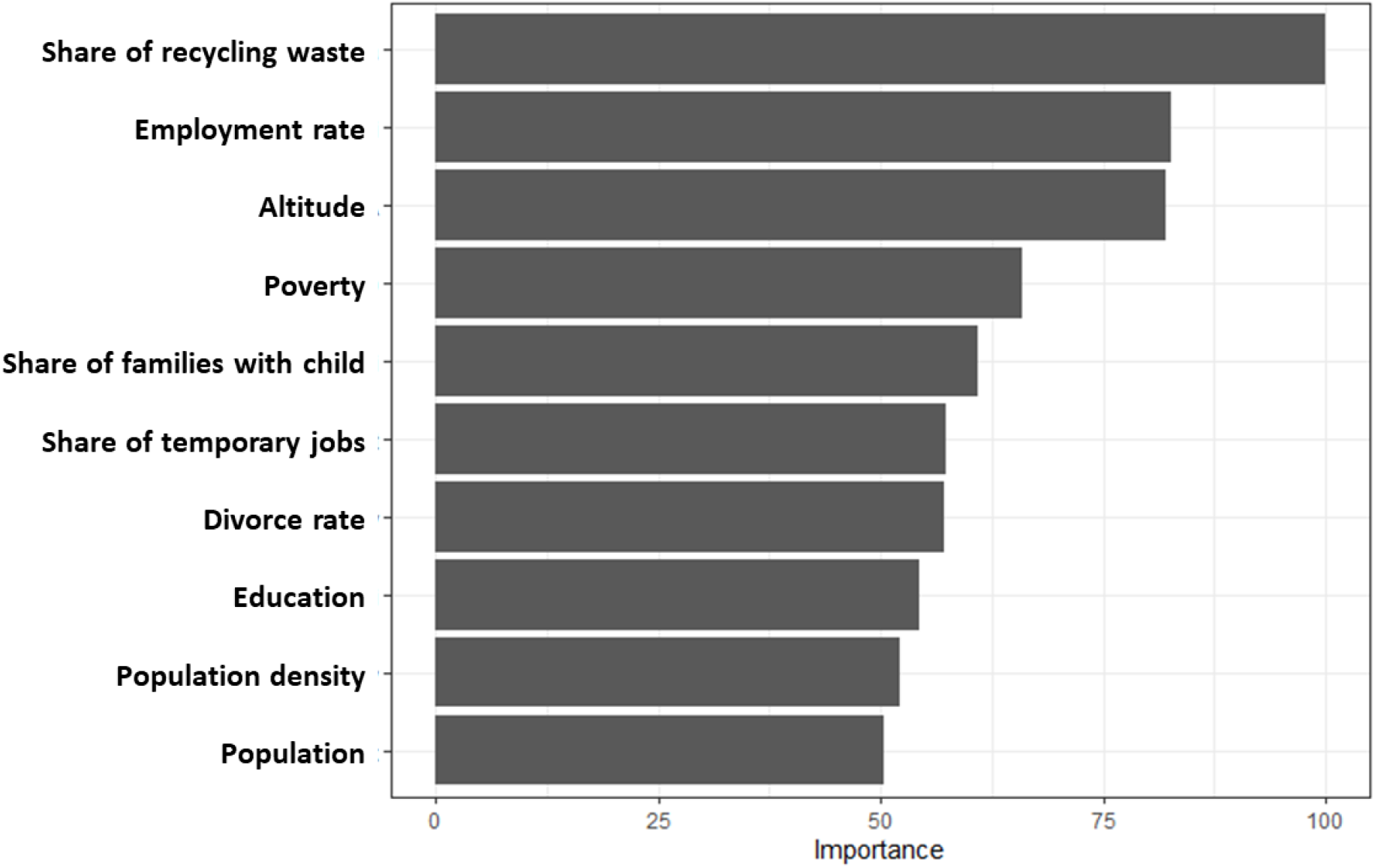
Feature Importance to predict municipal Vaccination Hesitancy for the first 10 important features in Random Forest. Random Forest trained on 70% of observations and tested on the remaining 30%; Resampling: Cross-Validated (10-fold, repeated 5 times. Hyper-parameters Best Tune for Random Forest is **mtry** (Number of variables available for splitting at each tree node) = 15.

The complete set of feature importance for the four models is shown in Table 4. A high and significant correlation (0.874; 95% CI = 0.743, 0.940) can be noted between the feature importance obtained by RF and GBM (the second-best performing model), while there is no significant correlation between the feature importance obtained between RF and the two worst performing models (LASSO and NN).

**Table 4.** Feature Importance for different Machine Learning Models

|  | RF | GBM | LASSO | NN |
| --- | --- | --- | --- | --- |
| Share of recycling waste | 100.00 | 100.00 | 13.89 | 44.52 |
| Employment rate | 82.73 | 42.26 | 17.64 | 16.05 |
| Altitude | 82.09 | 69.82 | 3.75 | 30.04 |
| Poverty | 65.84 | 63.61 | 46.36 | 12.22 |
| Share of families with Child | 60.97 | 34.71 | 8.22 | 8.28 |
| Share of temporary jobs | 57.39 | 29.16 | 11.13 | 19.73 |
| divorce rate | 57.17 | 32.01 | 8.20 | 9.62 |
| Education | 54.29 | 34.77 | 9.85 | 17.25 |
| Population Density | 52.12 | 21.91 | 1.22 | 14.68 |
| Population | 50.42 | 29.03 | 0.00 | 36.83 |
| Average Income | 43.42 | 35.07 | 0.00 | 100.00 |
| Average age of the council's members | 42.23 | 13.11 | 0.19 | 60.22 |
| Inequality | 41.35 | 22.65 | 100.00 | 2.07 |
| Share of population under 18 yo | 39.92 | 9.07 | 9.75 | 6.59 |
| share of population under 2 yo | 39.43 | 12.93 | 21.37 | 0.00 |
| share of population over 65 yo | 38.79 | 13.12 | 20.23 | 20.13 |
| Age of the Mayor | 34.34 | 6.99 | 0.01 | 45.42 |
| Share of females in the Council | 33.04 | 8.72 | 0.00 | 5.49 |
| Access to Internet | 25.96 | 24.33 | 3.38 | 26.85 |
| Special Statute Regions | 24.12 | 23.00 | 8.91 | 7.43 |
| Southern Region | 7.83 | 11.80 | 9.74 | 23.77 |
| Internal Area | 4.92 | 0.00 | 0.90 | 6.33 |
| Mayor with the degree or more | 4.57 | 1.66 | 2.17 | 10.47 |
| Earthquake | 4.50 | 9.99 | 2.57 | 27.81 |
| Mayor of Centre-Left | 4.42 | 0.00 | 1.81 | 11.66 |
| Mayor of Centre-Right | 4.02 | 0.95 | 3.03 | 14.45 |
| Female Mayor | 3.50 | 0.00 | 0.27 | 14.83 |
| Municipal Default | 0.00 | 0.00 | 8.38 | 1.65 |

In Figure 3 we finally show the Italian municipalities classified on the basis of VH risk. On the left panel, we report classification on the basis of real vaccination coverage data, while on the right panel we report the classification based on the predictions of our algorithm using lagged values of the area-level indicators (i.e. in 2015). A comparison of the two maps reveals the high potential of ML algorithm to predict VH risk. Figure 3 also shows a large territorial heterogeneity, with a higher VH risk found in rural areas far from metropolitan cities and in almost all the Southern regions and in particular in Puglia, Calabria and Sicilia.

**Figure 3.**
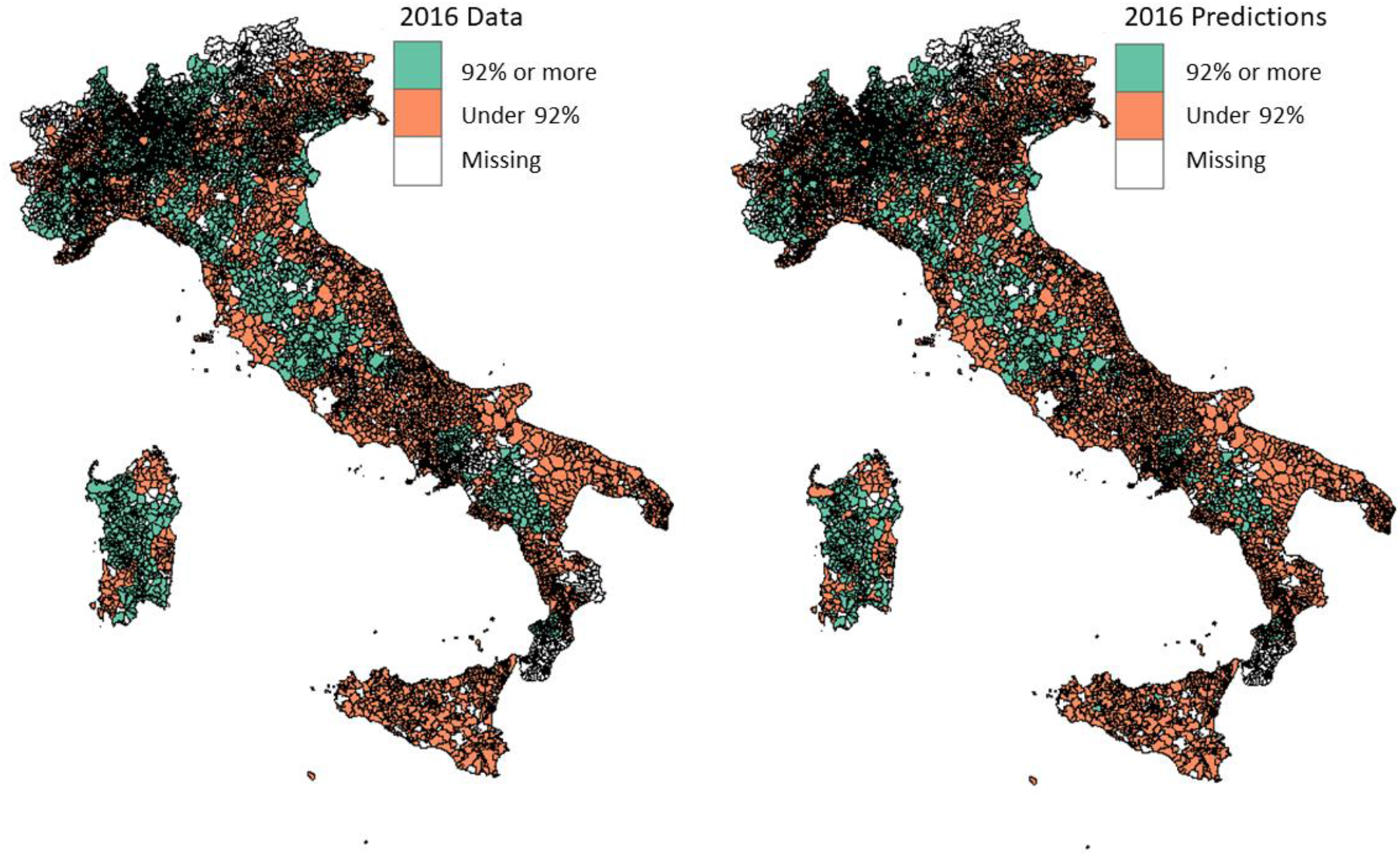
Vaccine Coverage Data (Left panel) and Predictions (Right panel) Random Forest trained on 70% of observations and tested on the remaining 30%; Resampling: Cross-Validated (10-fold, repeated 5 times. Hyper-parameters Best Tune for Random Forest is **mtry** (Number of variables available for splitting at each tree node) = 15.

## 4. Conclusions

This paper proposes a machine-learning approach for the prediction of vaccination hesitancy (VH) risk in an immunization campaign on the basis of area-level administrative data. We illustrate our approach using real-life data from child immunization campaigns done in Italy in 2016 concerning seven vaccine-preventable diseases. We find that the Random Forest algorithm is the best model to make out-of-sample predictions with a high true positive rate and a low false-positive rate. Compared with the unpredictable baseline level, the accuracy with Random Forest improves by 24%. A higher VH risk is found in rural areas far from metropolitan cities and in almost all the Southern regions and in particular in Puglia, Calabria and Sicilia. Moreover, we also find that the level of waste recycling and employment rate are the most important area-level predictors of a high VH risk in a community. The high association between VH risk and the level of waste recycling supports the relevance of pro-social behaviours in immunization choices (Crociata et al, 2015; Ramkissoon, 2020).

Although our analysis concerned child immunization, it is possible, albeit with the due limitations, to generalize our evidence to better face the mass immunization campaign for COVID-19. In fact, our approach using only lagged administrative data and socio-economic indicators at the municipal level is easy to be implemented in a short time and can be adapted to any vaccination campaign. Never as in COVID-19 vaccination campaigns, time is a crucial factor. Our approach thus offers a practical benchmark that can be used also in other countries to identify areas where the next mass immunization campaign for COVID-19 might experience low acceptance rates. This might allow policymakers to plan area-level pro-vaccine awareness campaigns.

To the best of our knowledge, the use of administrative data for learning and research purposes in vaccine campaigns is a relatively unexplored field. The main strength of administrative data is their immediate availability, at zero additional cost for analysts. Our approach also suggests that this does not come at costs in terms of accuracy, being the performances of our model in line with models using patient-level indicators to predict VH (see Bell et al. 2019, Oreskovic et al. 2020). The administrative data already exist and does not require an ad-hoc collection. The potential of such data is even greater in fragile settings where collecting (individual) data per se often presents several difficulties and there is pressure to act quickly.

On 19 January 2021, the European Commission [adopted](https://ec.europa.eu/commission/presscorner/detail/en/ip_21_143) a [Communication](https://ec.europa.eu/info/files/communication-united-front-beat-covid-19_en) calling on the Member States to speed up the roll out of vaccines across the EU. By March 2021, at least 80% of people over the age of 80, and 80% of health and social care professionals in every Member State should get vaccinated. By summer 2021, Member States should have vaccinated a minimum of 70% of the entire adult population. In order to achieve these goals, Health officials have to build trust in their communities through clear and transparent communication about vaccines. This includes information about their effectiveness, any expected side effects, and when to return for booster shots. Experts have raised alarm about increasing vaccine skepticism, which has already led to vaccine-preventable diseases outbreaks in several countries (Rosenthal 2010; Motta et al. 2018; Vinck et al. 2019). To prevent these events, it is necessary to act quickly and targeted campaigns using local indicators can certainly favour vaccination and prevent any hesitations.

## Data Availability

Raw data available upon request

## Footnotes

1 In March 2017, the so-called “Lorenzin law” was introduced to implement compulsory vaccination for children six years old and younger to be allowed into the school system.

2 In Italy the vaccination coverage has been included in the set of Essential Levels of Health Service (“*Livelli Essenziali di Assistenza*”, LEA). This set of services is defined at national level and provided by the public sector in each region. The recommended acceptable target of vaccination coverage for child immunization has been set to 95%. Below the 92% threshold, the situation is defined as “worrying”, and the regions must take action to increase vaccination coverage to avoid sanctions by the National Government.

